# AcuKG: a comprehensive knowledge graph for medical acupuncture

**DOI:** 10.1101/2025.03.22.25324466

**Authors:** Yiming Li, Xueqing Peng, Suyuan Peng, Jianfu Li, Donghong Pei, Qin Zhang, Yan Hu, Fang Li, Li Zhou, Cui Tao, Hua Xu, Na Hong

## Abstract

This study constructs an acupuncture knowledge graph (AcuKG) to systematically organize and represent acupuncture-related knowledge in a structured and scalable format. By extracting and integrating knowledge from diverse data sources, covering indication, treatment efficacy, practice guidelines, clinical research etc., AcuKG enhances knowledge discovery and utilization while improving data interoperability in the field of acupuncture. To achieve this, we employ multiple methods, including entity recognition, term normalization, semantic relation extraction, ontology mapping, etc., to extract and organize the acupuncture-related knowledge. Beyond knowledge structuring, to demonstrate AcuKG’s usability, we developed two use cases. First, AcuKG can assist obesity acupuncture knowledge discovery by linking key acupoints to obesity related acupuncture treatment efficacy research. Additionally, we demonstrate that knowledge injection into large language models (LLMs), such as ChatGPT and LLaMA, significantly improves their ability to answer acupuncture-related questions, increasing response accuracy. This work establishes a structured foundation for acupuncture knowledge representation, contributing to more reliable and efficient knowledge retrieval and discovery, and benefiting researchers, clinicians, and artificial intelligence (AI) applications in the field.

## Introduction

Acupuncture, a traditional Chinese medicine practice, has been widely accepted worldwide [1], [2], [3], [4], [5]. The number of licensed acupuncturists in the U.S. increased by 257% from 1998 to 2018, reaching a total of 37,886 [6]. Recognized by the National Institutes of Health (NIH) as a therapeutic intervention in complementary medicine ^5^, acupuncture is now primarily administered as an adjunctive treatment and complementary medicine [5], [6], [7, pp. 2002–2012], [8]. To alleviate various health conditions, acupuncturists use fine needles to insert into specific points on the human body, known as acupuncture points or acupoints [9]. According to ancient meridian theory, acupoints are located along meridians, which are channels forming a network system on the human skin where “Qi” (energy) flows [10]. Acupuncture helps unblock the Qi, allowing it to pass through the meridians and thereby treat diseases [10], [11]. Interest in acupoints has grown among physicians and researchers, with studies reporting their efficacy in treating conditions such as migraine, chronic pain, chemotherapy-induced nausea and vomiting, cancer, etc [1], [9], [12], [13], [14], [15], [16], [17], [18].

As acupuncture continues to gain recognition as a complementary therapy, there is a growing need to substantiate its efficacy with robust scientific evidence or explore the new potential efficacy of acupuncture. However, the traditional knowledge of acupuncture, often rooted in ancient practices, presents a unique challenge when it comes to integrating it with modern biomedical research and clinical data analysis [19], [20], [21]. One of the main reasons that impedes the efficient utilization of acupuncture knowledge for research and clinical application is that acupuncture knowledge suffers from fragmented and unstructured text spread across various sources, including books, web resources, traditional practices, research literature, and clinical trials[22]. The elements of acupuncture, such as acupoints, body locations, and measurement units, are plagued by inconsistent names, abbreviations, and coding systems across various studies [23], [24], [25]. Consequently, the data value and the translational value for acupuncture-related research, and evidence-based clinical practices are somewhat hindered.

Knowledge graph, which can provide a structured set of entities and their relations within a specific domain, facilitate data standardization, integration, knowledge representation, sharing, analysis, and mining [26], brings new potential for comprehensive analysis and practical application of acupuncture data. Previous efforts to build acupoint ontologies and knowledge graphs exist. For instance, Guan and Xie supplemented and adjusted the semantic types of acupoints and acupuncture methods, and designed a classification framework to construct acupuncture terminology sets [27]. Han et al. proposed the Acupuncture and Tuina Knowledge Graph to better understand and utilize knowledge from Traditional Chinese Medicine’s ancient literature [28]. Li et al. established a knowledge graph of acupuncture and tuina, including locations and indications, based on Chinese literature and web sources [29]. Existing standards such as Systematized Medical Nomenclature for Medicine–Clinical Terminology (SNOMED CT) and Read Codes (RCD) include acupoints but with limited information [30], [31]. While previous efforts have laid the foundation for acupuncture knowledge representation, they remain limited in scope, they have often been coarse-grained, lacking detailed semantics, aggregated knowledge, or clinical evidence [27], [28]. Therefore, establishing a data-driven, evidence-based, and computationally structured acupuncture knowledge system that can align with modern clinical practices is crucial but remains an unresolved challenge.

To address these challenges and bridge the gap between clinical practice, scientific research, and digital applications in the field of acupuncture, this study aims to capture the intricate details of acupuncture points, their clinical indications, and associated clinical evidence to construct a comprehensive, unified and structured repository of acupuncture knowledge. Previously, we have extracted the acupoint-related entities (acupoint, anatomy, direction, distance, general location, and subpart) and relations (’direction_of’, ‘distance_of’, ‘part_of’, ‘near_acupoint’, and ‘located_near’) from the World Health Organization Standard Acupuncture Point Locations in the Western Pacific Region (WHO Standard) with machine learning models and large language models respectively [10], [32]. Building on our previous work, we expanded beyond acupuncture location-related knowledge extraction from WHO Standards to integrate a broader range of acupuncture-related information from diverse sources, creating a comprehensive knowledge graph that provides a multidimensional perspective on acupuncture knowledge.

This study aims to develop a structured and comprehensive knowledge graph that integrates fragmented acupuncture knowledge and facilitates its application in acupuncture research and clinical practice. By consolidating fragmented data from multiple high-quality resources, including acupuncture online resources, literature, clinical trials, and specialized ontologies, we curated AcuKG, a knowledge graph designed to encapsulate a wide range of data while providing comprehensive, standardized, and integrated acupuncture knowledge.

## Material and Methods

In this study, we utilized various acupuncture-specific data sources to collect, organize, and represent acupuncture-related knowledge. As shown in Figure 1, this section provides a detailed overview of the data sources used, the processes of entity extraction, relation extraction, the ontology mapping approach, and the graphical representation of our knowledge graph AcuKG. Furthermore, we present two use cases designed to evaluate the usability and effectiveness of the AcuKG.

**Figure 1.**
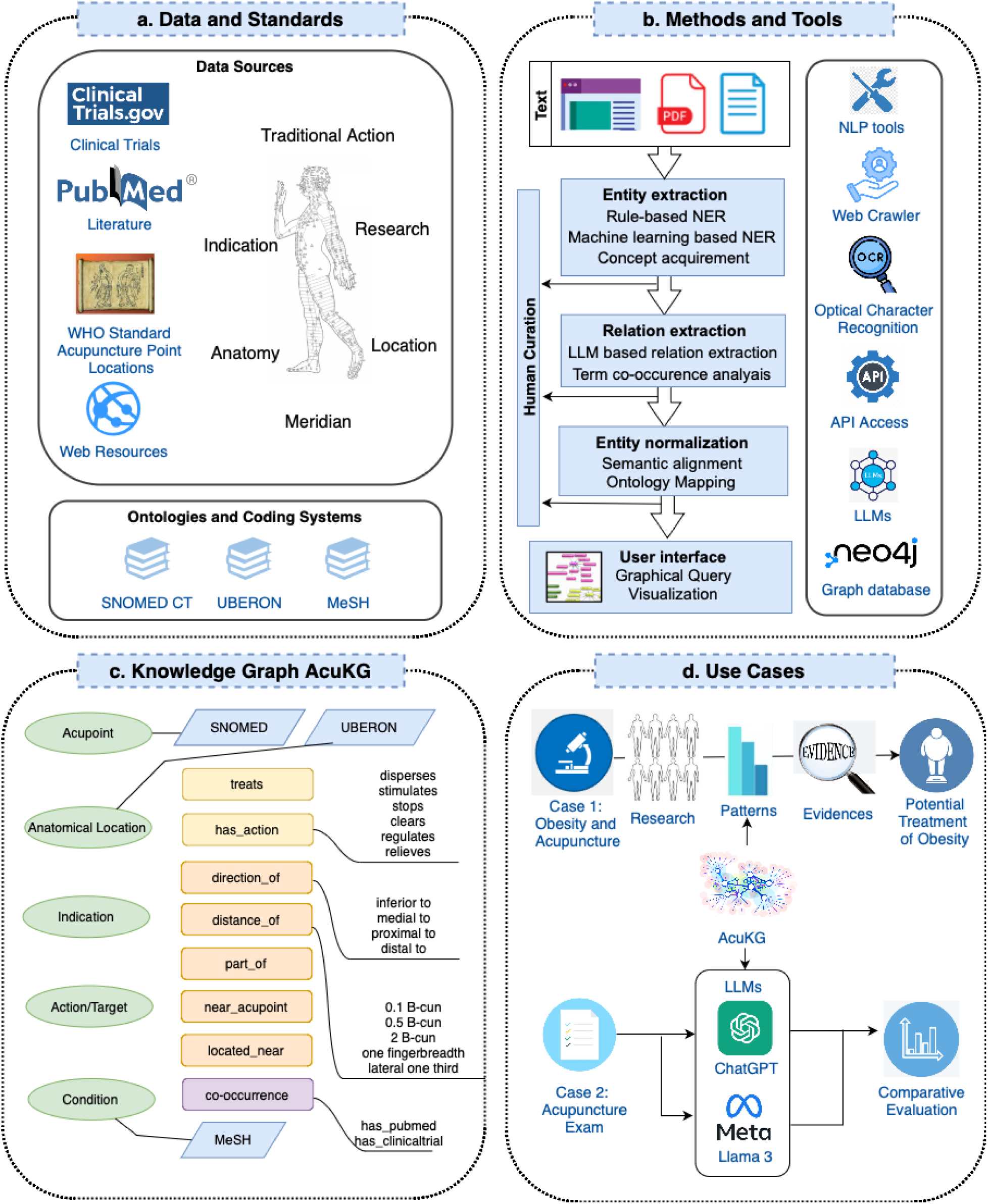
Overall framework of the study.This figure illustrates the structured workflow of AcuKG construction and application. (a) **Data and Standards**. Various data sources, including clinical trials, biomedical literature, WHO standard, and web resources with ontologies and coding systems like SNOMED CT, UBERON, and MeSH**. (**b**) Methods and Tools**. Entity and relation extraction, ontology mapping, and natural language processing techniques were employed to build the knowledge graph. (c) **Knowledge Graph AcuKG**. A structured representation capturing relationships between acupoints, indications, anatomical locations, and action targets. d(d**) Use cases**. Demonstrates AcuKG’s applicability in clinical decision-making and knowledge discovery.

### Data sources

The data sources used in this study included a diverse range of credible databases and resources to ensure comprehensive acupuncture knowledge coverage and quality. We gathered acupuncture knowledge from WHO Standard, specialized acupuncture web resources, clinical trial registry, and clinical study articles indexed by PubMed (Table 1) [33], [34], [35]. Additionally, to ensure standardized terminology and interoperability in our knowledge graph, we linked our knowledge graph to multiple ontologies and coding systems that are well established in the field of biomedical and clinical research (Table 1), such as UBERON, SNOMED CT, and Medical Subject Headings (MeSH) [36], [37], [38], [39]. These data sources provided a robust foundation for building a fine-grained and reliable knowledge graph, facilitating the standardization and integration of acupuncture-related information.

**Table 1.** Description of data sources.

| Type of data source | Data source | Data format | Collected Data |
| --- | --- | --- | --- |
| Acupuncture knowledge | WHO Standard [33] | PDF | Acupuncture points, and description about the acupoint location |
|  | Acupuncture Web Resources [34] | HTML | Acupuncture points, Indications, Actions |
|  | Clinicaltrial.org [35] | JSON | Acupuncture points, Conditions |
|  | PubMed | XML/PDF | Acupuncture points, Conditions |
| Ontologies/Coding systems | SNOMED CT [40] |  | Indications, Actions, Anatomical locations, and Acupuncture points |
|  | MeSH [39] | JSON | Conditions |
|  | UBERON [37] | CSV | Anatomical locations |

### Acupuncture Knowledge Sources

(1) *WHO Standard* was developed to harmonize acupuncture practices by providing consistent, scientifically validated references for 361 point locations [33]. Developed through expert collaboration and comprehensive reviews, It serves as an essential resource for practitioners, educators, and researchers, facilitating the integration of acupuncture into modern healthcare systems and supporting international collaboration [33]. (2) *Acufinder.com* is a comprehensive online resource dedicated to acupuncture and traditional oriental medicine [34], [41], [42], [43]. The website offers detailed information on each acupuncture point, including its code, Chinese name, English name, location, indications, and traditional actions. By providing this extensive knowledge, Acufinder.com aims to support the professional growth of acupuncturists and facilitate research in the field of acupuncture and oriental medicine. (3) *PubMed* is a free, publicly accessible database maintained by the U.S. National Library of Medicine (NLM) that provides comprehensive coverage of biomedical and life sciences literature [44]. We retrieved all literature related to acupoints from PubMed, focusing on clinical studies [45]. Our MeSH term based query ensured that the retrieved literature was relevant, of high quality, and reflected the most up-to-date knowledge in acupuncture research. (4) *ClinicalTrials.gov* is a publicly accessible database maintained by the NLM that provides information on clinical studies conducted worldwide [35]. We conducted a systematic search of ClinicalTrials.gov using a predefined acupuncture point vocabulary [45] to identify relevant clinical trials involving acupuncture.

### Ontologies and Coding Systems

(1) Uber anatomy ontology (UBERON) is an integrated cross-species anatomy ontology representing various entities classified based on traditional anatomical criteria like structure, function, and developmental lineage [37]. (2) SNOMED-CT is the most comprehensive, multilingual clinical healthcare terminology that provides codes, terms, definitions, synonyms, and definitions of clinical findings, symptoms, diagnoses, procedures, body structures, organisms and other etiologies, substances, pharmaceuticals, devices, and specimens used in the clinical documentation and reporting [38], [46], [47]. (3) MeSH is the controlled vocabulary used to index journal articles and books in the biomedical domain, especially PubMed [48]. MeSH is also referenced by the ClinicalTrials.gov registry to classify the diseases studied by the clinical trials [48].

### Building the Acupuncture Knowledge Graph

To construct our comprehensive knowledge graph, We first defined the scope of five types of entities and eight types of relationships to be extracted from various sources. The entities include acupoint, action target, indication, condition and anatomical location, while the relationships encompass *direction_of*, *part_of*, *distance_of*, *near_acupoint*, *located_near*, *has_target*, *treats*, and *co-occurrence*. Then, we undertook meticulous data processing and extraction steps for diverse data sources, including OCR, entity recognition, term normalization, semantic relation extraction, ontology mapping, etc. The specific methods and data organization process are outlined below.

### Entity extraction

#### (1) Optical Character Recognition (OCR)

The WHO Standard is provided as a searchable PDF, and OCR models were used to extract key information embedded in this document. We extracted acupoint codes, names, body parts, and location descriptions, converting them into XML format. Our approach utilized PDF content identification techniques based on the Apache PDFBox package, which efficiently facilitates text extraction from WHO Standard PDF document [49]. Our method employed a two-step process: (1) PDF layout analysis to identify logical sections (e.g., header, footer, abstract) based on page numbers and column order, and (2) graphic coordinate analysis to pinpoint tables, figures, and images within each block. Each acupoint section is structured with text descriptions on the left and corresponding images on the right, separated by lines, this standardized layout ensures precision and efficiency in extraction.

#### (2) Rule based NER

For the web-based data from Acufinder.com, we employed rule-based natural language processing (NLP) methods to extract indications and actions [50]. This process involved splitting sentences, removing stop words, and restructuring phrases to standardize the concepts. Indications, represented as noun phrases or prepositional phrases, were extracted and processed accordingly. Noun phrases were obtained by splitting elements separated by commas, while prepositional phrases were handled by generating combinations of nouns and verbs before and after the preposition if only one preposition and one conjunction were present. For example, the indication phrase “pain in the elbows and arm” was restructured into “elbows pain” and “arm pain.” In cases with more complex structures, such as “redness and swelling of fingers and the dorsum of the hand,” manual splitting was performed. For actions, verb-object collocations were extracted by removing stop words (e.g., “a,” “an,” “the”), and then splitting phrases based on punctuation (commas, periods) and conjunctions (e.g., “and,” “or”) to ensure consistent representation.

#### (3) Machine learning based NER

For the acupoint location entity extraction from the text description within WHO Standard, we used multiple machine learning based NLP methods, including Conditional Random Fields (CRFs) and Recurrent Neural Networks (RNNs) to extract entities like *Anatomy*, *Direction*, *Distance*, *General Location*, and *Subpart* [10]. CRFs were applied for sequence labeling tasks to extract entities using n-grams and word embeddings as features. RNNs were employed to capture sequential dependencies and identify relationships between acupoints and anatomical locations. The results were evaluated using 5-fold cross-validation on annotated datasets, achieving high accuracy in extracting acupoint-related entities. These methods were comprehensively described and validated in our previous study [10]. In this study, we integrated the entity extraction results by the best performed models to the AcuKG knowledge graph.

#### (4) Concept extraction based on terminologies

For clinical trials, we used the application programming interface (API) of clinicaltrials.org to collect clinical trials that contain acupuncture points based on our predefined acupoint vocabulary. For each clinical trial, all the acupuncture points information, and all the relevant MeSH terms were extracted, which represents the primary disease or condition being studied in the trial. For the PubMed literature, we conducted several steps to extract the entities of acupuncture points and conditions. First, we obtained all the full-text articles available in PubMed Central (open access dataset) from the acupuncture literature we retrieved. Then, we extracted the methods and results sections of each article, as these are most likely to contain the research questions and findings related to acupuncture points and their associated conditions [51]. Next, for each MeSH-indexed article from PubMed, we extracted all their MeSH terms and filtered those categorized as representing the primary disease or condition using the same MeSH term subset as clinical trials [45]. Subsequently, we also used our predefined acupoint vocabulary [45] that contains all the acupoint codes and names, to capture the Acupoint terms from each article. This concept extraction method allowed us to focus on the identification of acupuncture points and conditions from the literature.

### Relation extraction

#### (1) LLM based relation extraction

To build the location relations of acupuncture points, we included relations annotated in our previous study, which was based on acupoint descriptions from the WHO Standard [32]. Five types of location relations are defined to accurately represent acupoint locations: *direction_of*, indicating the direction of one relative acupoint or anatomy to the acupoint of interest; *distance_of*, denoting the distance to the acupoint or anatomy; *part_of*, representing the subpart of the anatomy; *near_acupoint*, referring to adjacency to the relative acupoint; and *located_near*, signifying the proximity of the anatomy to the acupoint of interest. Multiple LLM based relation extraction methods were explored and validated in our previous study [32]. In this study, we integrated the relation extraction results to the AcuKG knowledge graph.

#### (2) Terms co-occurrence analysis

Based on the extracted entities of acupuncture points and conditions from literature and clinical trials, condition terms were paired with the acupoints in the same literature or clinical trial using the co-occurrence analysis method. In terms of different data sources, we specify the *co-occurrence* relations into two subtypes of co-occurrence relations: *has_pubmed* and *has_clinicaltrials*. These pairs, along with their National Clinical Trial Identifier Numbers (NCT IDs) or PubMed reference numbers (PMIDs), provided a comprehensive and fundamental structured network for building acupoint-condition relations for the acupuncture knowledge graph.

### Entity normalization and ontology mapping

For the Acufinder.com online data, two authors reviewed extracted acupoints names, indications, and actions associated with each acupoints[50]. Inconsistent names and expressions were manually corrected and normalized to the accurate term name. To further enhance entity standardization level, we performed name matching of acupoints and anatomical location terms with concepts and their corresponding synonyms from SNOMED CT and UBERON. This ensured the use of consistent terminology and facilitated the accurate semantic representation of data in our knowledge graph. This structured approach allowed seamless integration of extracted acupuncture knowledge into our comprehensive knowledge graph AcuKG. Additionally, we linked each PubMed article’s PMID and each Clinical Trial’s NCTID for quick indexing.

### Graph query and visualization

AcuKG was visualized using Neo4j software with py2neo Python package, a client library and toolkit for working with Neo4j from within Python applications and from the command line [52], [53]. The data were first transformed to a triple format suitable for Neo4j with properties/attributes attached to entities and relations. Entities such as specific *Acupoint*, *Anatomical location*, and *Condition* were represented as nodes, while relations such as *treats* and *located_near* were defined as edges. In addition, we added the Chinese names, pinyin names, and MeSH IDs as properties/attributes of acupoints and MeSH terms. The NCTIDs and PMIDs were added as properties/attributes of relations from clinical trials and literature. Neo4j’s Cypher query language was used for searching and querying. Overall, with the graphical interface, AcuKG allows clinicians and researchers to visualize data and facilitates easier knowledge retrieval and querying.

### Evaluation Methods

To ensure the content accuracy of AcuKG, we implemented the following processes: (1) two authors conducted a manual review of the extracted content from the acupuncture web resource and the WHO Standard; and (2) for the content extracted from PubMed and Clinical Trials, entities were extracted based on predefined vocabularies or annotated terms, so they are normalized and consistent. Meanwhile, to maximize the accuracy of the relations, we applied a frequency-based filtering approach, single-occurrence relations were excluded to reduce error relations that may result from extraction errors or occasional mentions. Only acupoint-condition pairs that appeared at least twice were included

To evaluate the usability of AcuKG, we designed two use cases. The first use case focuses on obesity and acupuncture knowledge discovery, given that obesity is a global health concern associated with chronic diseases, while acupuncture shows potential as a complementary treatment. However, the relationship between acupuncture and obesity, as well as its efficacy, remains underexplored. Meanwhile, large language models (LLMs) show promise in medical applications but lack accuracy in specialized domains like acupuncture. In the second use case, we tested whether GPT-4o and LLaMA 3 improved their accuracy in acupuncture-related question answering by incorporating AcuKG as structured input data, measuring correctness via a real acupuncture exam. These use cases demonstrated AcuKG’s application scenarios and the capability of empowering AI applications in the Acupuncture domain.

## Results

### Entities and relations in AcuKG

AcuKG was constructed with triples as entities and relations between entities. A total of 1,839 entities were created, encompassing five types in our KG(Figure 2 (a)): *acupoint* (361, 19.63%), *action target* (230, 12.51%), *indication* (447, 24.31%), *anatomical location* (404, 21.97%), and *condition* (397, 21.59%). Every acupoint has its English name, Chinese name and Chinese pinyin name as properties/attributes. Every MeSH term has its corresponding MeSH ID as properties/attributes.

**Figure 2.**
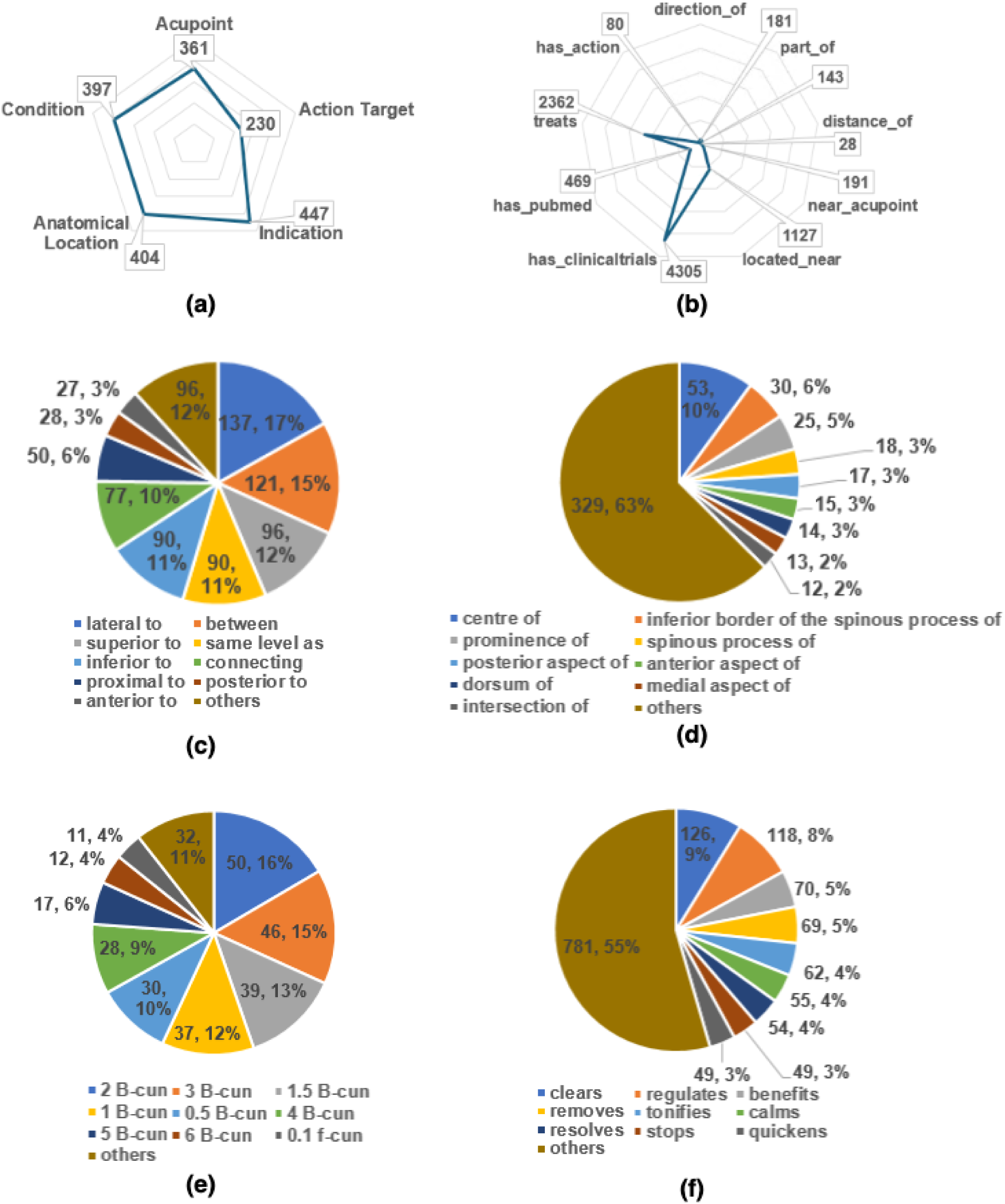
Distribution of (a) **entities**: Five types of entities, including *acupoint*, *indication*, *action target*, and *anatomical location*, are represented in the knowledge graph; (b) **relations**: Various relation types and their frequencies, such as *direction_of*, *part_of*, *treats*, etc. Subtype of (c) **direction_of**: Common spatial relations include “lateral to” (17%) and “between” (15%), with other directional terms distributed accordingly; (d) **part_of**: Hierarchical anatomical structures are represented, with “centre of” (10%) and “inferior border of the spinous process of” (6%) being the most frequent; (e) **distance_of**: Key spatial measurements, including 2 B-cun (16%) and 3 B-cun (15%), are identified; (f) **has_action**: Functional relations of acupoints are highlighted, with “clears” (9%) and “regulates” (8%) as the most prevalent therapeutic actions.

The KG also defines various semantic relations between these entities. Among the 11,527 established relations, an overview of their types and frequencies in the KG is presented as follows (Figure 2 (b)): *direction_of* (812, 7.04%), *part_of* (526, 4.56%), *distance_of* (302, 2.62%), *near_acupoint* (191, 1.66%), *located_near* (1127, 9.78%), *has_clinicaltrials* (4305, 37.35%), *has_pubmed* (469, 4.07%), *treats* (2362, 20.49%), and *has_action* (1433, 12.43%). For relations between acupoints with conditions, the NCT IDs of the clinical trials or PMIDs of the PubMed articles, were also collected as properties/attributes of the corresponding relations.

The spatial relation *direction_of* plays a critical role in acupuncture, as understanding these distributions allows for better modeling of acupoint placements and their potential interactions with physiological structures. Figure 2(c) shows that the most common subtypes of *direction_of* (n=39) are “lateral to” (17%) and “between” (15%), followed by “superior to,” “same level as,” and “inferior to” (each around 11-12%). “Connecting” (10%) and “proximal to” (6%) are moderately frequent, while “posterior to” and “anterior to” are less common (around 3% each). The remaining 12% fall into the “others” category.

The *part_of* relation is particularly important in defining hierarchical structures within acupuncture, as many acupoints are defined based on their position relative to larger anatomical locations. Figure 2 (d) illustrates the distribution of the *part_of* relation subtypes (n=143). The majority fall into the “centre of” (10%) and “inferior border of the spinous process of” (6%) category, with other subtypes having smaller proportions.

The *distance_of* relation is particularly relevant in acupuncture practice, as the precise measurement of acupoint spacing is crucial for accurate needling and treatment planning. Figure 2 (e) shows the distribution of the *distance_of* relation subtypes (n=28). The most common measurements are 2 B-cun (16%), 3 B-cun (15%), and 1.5 B-cun (13%). The “others” category accounts for 11% of the total.

The *has_action* relation describes the functional effects of acupoints on physiological processes, making it essential for understanding the therapeutic mechanisms of acupuncture. Finally, the distribution of the *has_action* relation subtypes (n=80) is illustrated in Figure 2 (f). The most prevalent actions are “clears” (9%) and “regulates” (8%), followed by “benefits,” “removes,” and “tonifies”, each accounting for 4-5% approximately.

### Mappings to external ontologies

Table 2 provides a breakdown of entities that are mapped to different external ontologies, along with the number of mappings for each entity type. It includes *acupoint* entities (n=361), *indication* entities (n=446), and *action target* entities (n=229) mapped to SNOMED CT. *anatomical space* (n=8) and *anatomical location* entities (n=395) mapped to UBERON, in addition to SNOMED CT. These mappings to external ontologies enhance the structured representation and interoperability of acupuncture-related concepts, facilitating their integration and utilization in various biomedical applications.

**Table 2.**
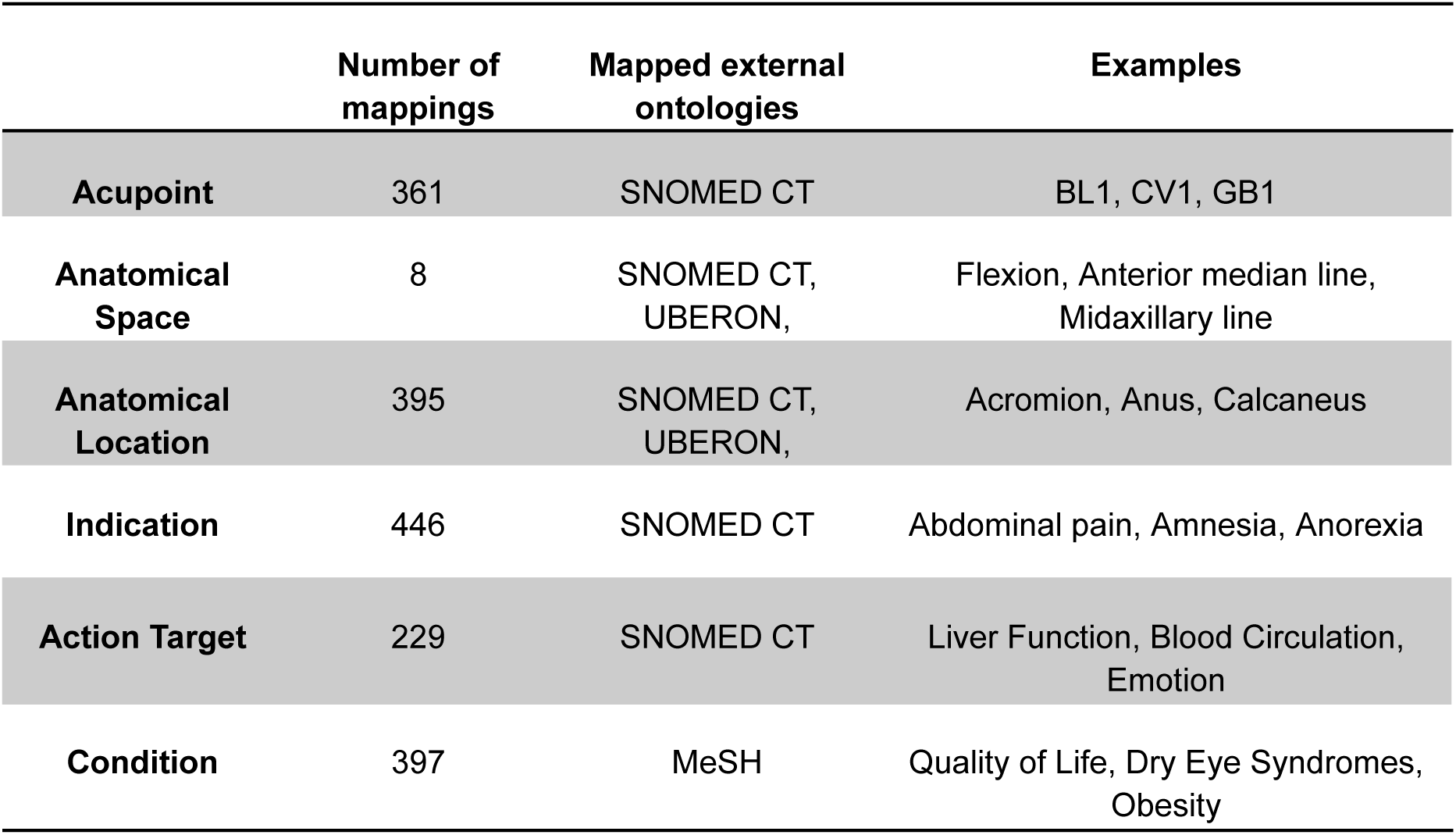
Overview of the mapped entities in the external ontology.

### Graphical query with visualized user interface

AcuKG provides retrieving and querying functions for users to better support our data application. Table 3 shows some example queries to simulate real application questions. Here, For example, a researcher may need to investigate all the information about one particular acupoint, like HT3. With query 1, we can get all the relations of acupoint “HT3”. In our result (Figure 3), HT3 related to 3 *action targets*, 9 *indications*, 3 *anatomical locations* and 43 MeSH terms including its names, locations, potential targets, and research information. With this organized information. With our KG, it would be easier for researchers to find the potential target diseases or symptoms for a particular acupoint and help them narrow down the research scope and find valuable research recommendations.

**Figure 3.**
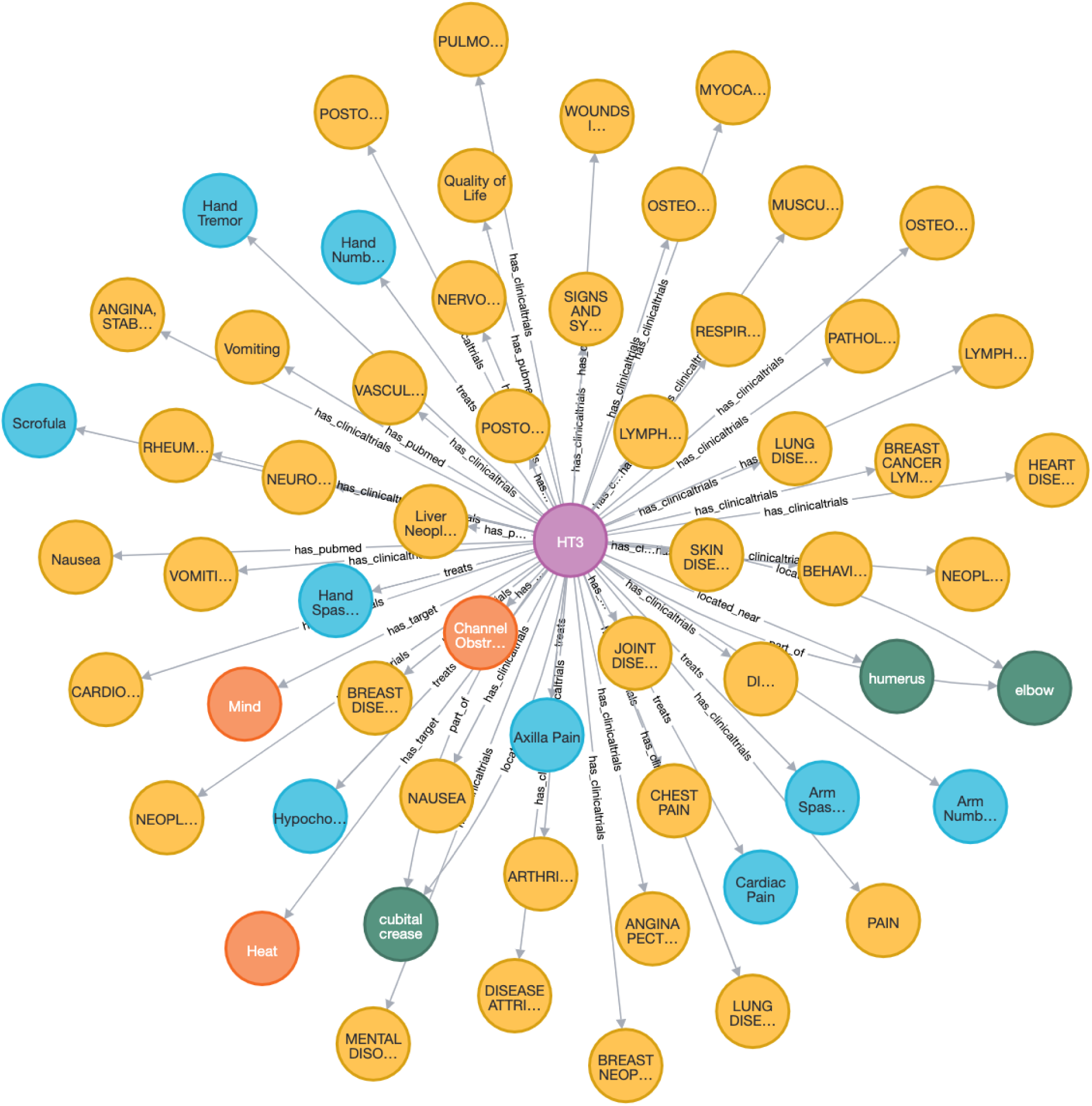
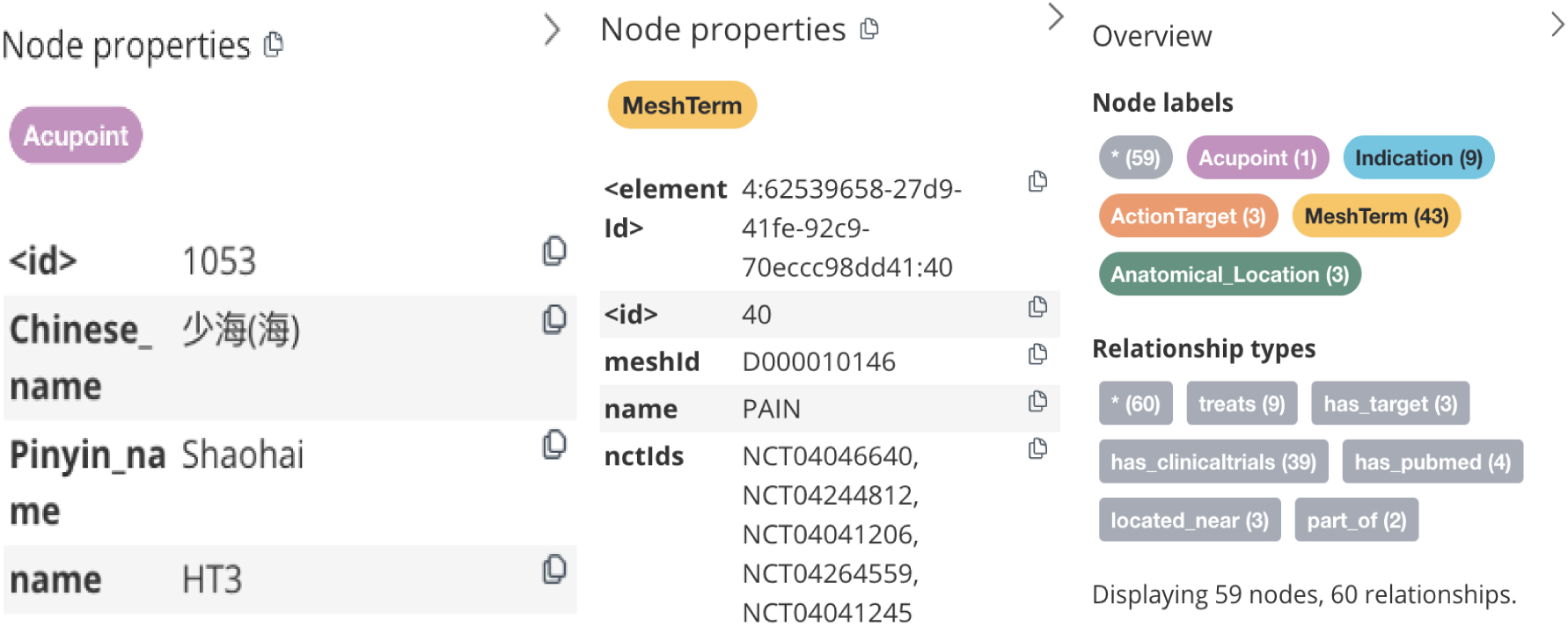
Graphical Query Results of Acupoint HT3 (Query. **Example 1**) This figure presents the query results for HT3, illustrating its associations with various entities in AcuKG. HT3 is linked to three action targets, nine indications, three anatomical locations, and forty-three MeSH terms.

**Table 3.** Example Queries.

| No | Questions in human language | Queries in Cypher query language |
| --- | --- | --- |
| 1 | What can I know about the acupoint “HT3”? | MATCH (n{name:"HT3"})--(o) RETURN n,o |
| 2 | Which acupoints should I choose for depression? | MATCH (n:Acupoint)--(o{value:"Depression"})<br>RETURN n,o |
| 3 | Where is the acupoint “HT3”? | MATCH (n:Acupoint)-[r]-(o{value:"Stomach"})<br>RETURN * |
| 4 | Which clinical trials and PubMed articles have investigated the potential relationship between HT3 and vomiting? | MATCH (n{name: "HT3"})--(o{value:"Vomiting"})<br>RETURN * |

### AcuKG Use Cases

#### Use Case 1: Accelerate obesity acupuncture knowledge discovery

Obesity is a global health issue characterized by excessive body fat accumulation that poses significant risks to health [54], [55]. According to the World Health Organization (WHO), the prevalence of obesity has tripled since 1975, with over 650 million adults worldwide classified as obese as of 2016 [56]. In the United States, the Centers for Disease Control and Prevention (CDC) reports that approximately 42.4% of adults were considered obese in 2017-2018 [57]. Obesity is associated with a range of comorbidities, including type 2 diabetes, cardiovascular diseases, and certain cancers, making it a critical target for public health interventions [58], [59].

Acupuncture has been utilized as a complementary therapy for obesity management, with several studies indicating its potential benefits in weight reduction and metabolic regulation [60]. To evaluate the utility of our acupuncture knowledge graph, we developed a use case focusing on identifying and understanding the acupoints related to obesity. This use case aims to map the relevant acupoints, their traditional functions, and their associations with clinical outcomes in obesity management, providing a structured conceptual framework to support further research and clinical applications in this domain.

AcuKG can facilitate obesity knowledge discovery by linking various acupuncture knowledge relevant to obesity. Several acupoints have emerged as key areas of focus in both literature and clinical trials for managing obesity. ST25 stands out as the most extensively studied acupoint, with 15 clinical trials and 3 publications (PMIDs: 33235115, 27457720, 31775839). This indicates a strong foundation in both traditional and evidence-based research. ST36 follows closely, being cited in 14 clinical trials and 3 publications (PMIDs: 27457720, 33235115, 31775839), demonstrating its established role in obesity management. CV12 is another notable acupoint, highlighted in 13 clinical trials and 3 publications (PMIDs: 27457720, 31775839, 33235115), reflecting its widespread recognition in both domains.

Other acupoints also show moderate levels of attention across both literature and trials. For example, CV6 and GB26 are each mentioned in 7 clinical trials and 2 publications (PMIDs for CV6: 27457720, 31775839; PMIDs for GB26: 27457720, 31775839), while SP15 appears in 8 clinical trials and 2 publications (PMIDs: 27457720, 31775839). These points indicate a growing interest and consistent focus on validating their therapeutic relevance. ST40 also deserves attention, with 6 clinical trials and 3 publications (PMIDs: 33235115, 27457720, 31775839), suggesting a balance between theoretical acknowledgment and clinical exploration.

Other acupoints such as BL2, KI12, ST1, ST2, and ST3 are predominantly referenced in literature, often appearing in 2–4 publications but with limited representation in clinical trials. Conversely, acupoints like CV10, CV4, and ST28 are heavily studied in clinical trials, appearing in 3–6 trials each, despite their minimal mentions in the literature. This contrast highlights the evolving focus of research, with some acupoints moving from theoretical exploration to empirical validation, while others remain underrepresented in clinical settings.

This dual emphasis on theoretical understanding and empirical validation enriches the evidence base for acupuncture’s role in obesity management. Acupoints like ST25, ST36, and CV12 exemplify this integration, serving as central figures in both traditional and clinical research while emerging points such as CV10 and ST28 highlight new directions for future studies.

The knowledge graph (Figure 4) representing acupoint locations associated with obesity comprises 92 nodes, including 40 *acupoints* and 52 *anatomical location* entities. The graph captures a total of 225 relationships, categorized into 210 *direction_of* and 15 *near_acupoint* relationships. The overview highlights a structured organization of the graph, while the example illustrates specific relationships, such as the spatial connections of acupoints (e.g., ST1, ST2, ST3) with nearby anatomical features like the nose, pupil, and face. This structured representation enables precise spatial understanding of acupoints, facilitating integration into clinical and computational frameworks for obesity treatment and research.

**Figure 4.**
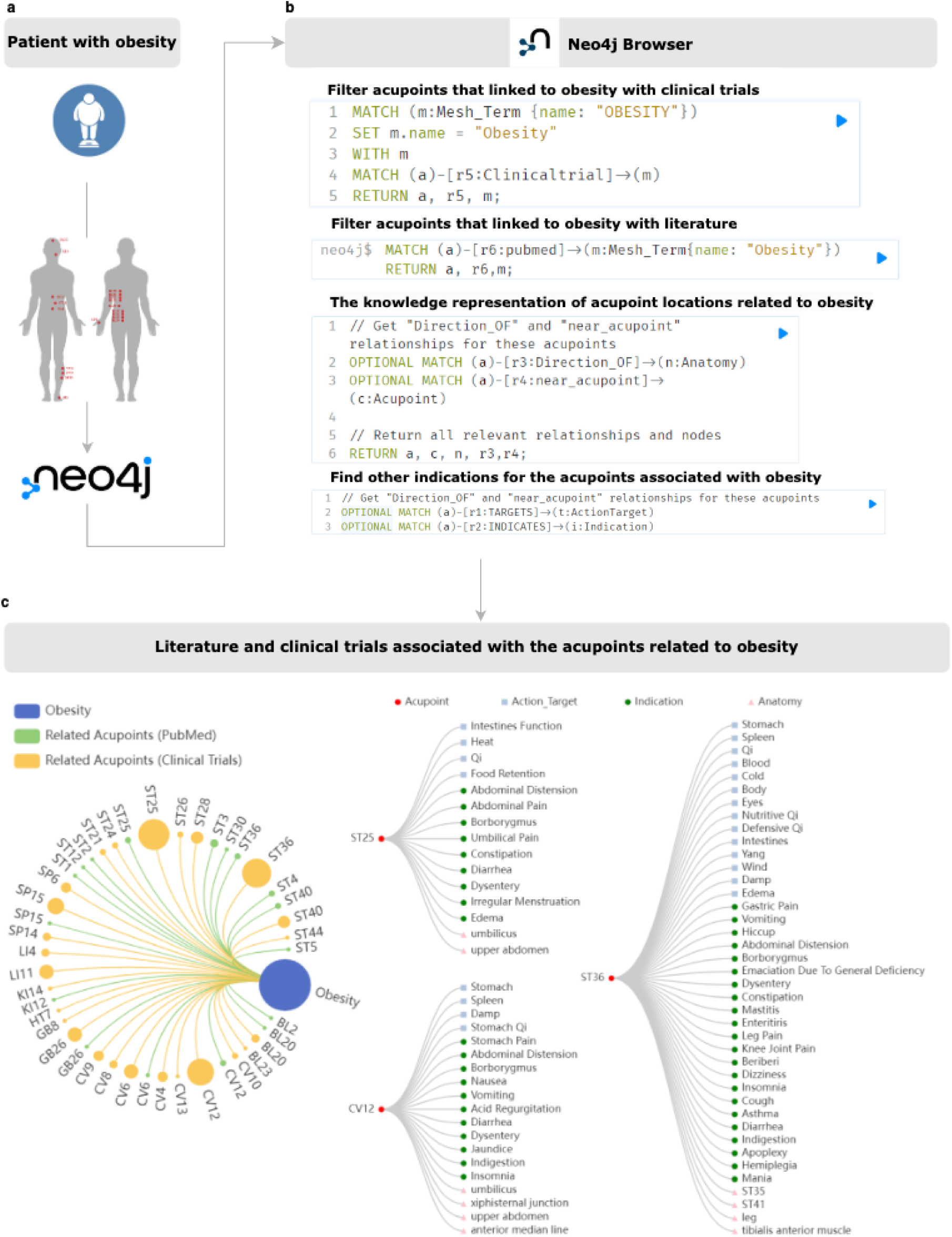
Overview of obesity acupuncture knowledge discovery. This figure illustrates the structured integration of acupuncture knowledge related to obesity (a) **Schematic overview**. Highlights AcuKG’s role in obesity management, mapping key acupoints and their relations with clinical outcomes (b**) Neo4j interface**. Demonstrates interactive query capabilities for exploring acupoint-obesity relations (c**) knowledge graph representation**. Presents 92 nodes (40 *acupoints*, 52 *anatomical location* entities) and 225 relationships (210 *direction_of*,15 *near_acupoint*), enabling spatial analysis of acupoints, which have strong clinical and literature support.

#### Use Case 2: Enhancing LLM acupuncture-related knowledge accuracy with the integration of AcuKG

In recent years, large language models like Generative Pre-trained Transformer (GPT) and LLaMA have garnered significant attention for their ability to generate human-like text across a variety of tasks [32], [61], [62], [63], [64], [65], [66], [67]. GPT, developed by OpenAI, has demonstrated remarkable capabilities in understanding and generating natural language, making it useful for a wide range of applications, including answering medical queries [68], [69], [70], [71], [72]. Notably, GPT-3 and GPT-4 have shown impressive performance in the United States Medical Licensing Examination (USMLE), achieving scores that are comparable to or even exceed those of passing medical students [73]. For instance, a study by Kung et al. reported that ChatGPT significantly performed at or near the passing threshold of 60% for all three exams without any specialized training or reinforcement [74]. This impressive performance has led to increasing interest in the potential applications of LLMs in medical education, where they could serve as tools for learning, assessment, and even clinical decision support.

Building on the success of GPT-4, OpenAI introduced GPT-4o, an optimized version designed to enhance accuracy, contextual understanding, and efficiency across more specialized and complex domains [75]. GPT-4o integrates advancements in model architecture and training techniques, allowing it to handle intricate queries with improved precision [76]. This makes GPT-4o particularly well-suited for applications requiring a deep understanding of specialized fields, such as medicine.

Similarly, LLaMA 3, developed by Meta, represents another significant advancement in the field of LLMs [32]. Known for its robust performance in various NLP tasks, LLaMA 3 incorporates improvements in model training and architecture, which enhance its capabilities in handling specialized knowledge and complex queries [32]. Its design focuses on achieving high accuracy and contextual relevance, making it a valuable tool for domain-specific applications as well.

Despite these advancements, the accuracy of GPT-4o and LLaMA 3 in specialized areas like acupuncture remains uncertain. Acupuncture, with its complex and nuanced body of knowledge, poses a particular challenge [10]. For instance, acupoint locations and their clinical indications are often tied to Traditional Chinese medicine (TCM) theories that may not be fully captured by a general-purpose LLM [32]. This discrepancy raises questions about the reliability of LLMs in providing accurate and contextually relevant information in specialized fields [32].

To explore the potential of enhancing the performance of ChatGPT powered by GPT-4o and LLaMA 3 in the domain of acupuncture, we conducted a use case study as shown in Figure 5. In this study, we imported acupuncture related questions from the Internal Medicine Question Bank for Chinese Medicine (2020, Acupuncture part) with the prompts related to acupoint locations and indications using both ChatGPT and LLaMA3. The questions covered a range of acupoints and their traditional uses, reflecting common clinical queries. Initially, we posed these questions directly to ChatGPT and LLaMA3 without any additional resources or domain-specific knowledge integration (prompts are provided in supplementary files). The model’s responses were evaluated based on their accuracy in providing correct and contextually appropriate information about acupoints. We further integrated our proposed AcuKG into the process. It begins with establishing a connection to a Neo4j-based KG containing acupuncture knowledge, clinical trials, and PubMed references. We extracted relevant data using structured queries to retrieve relations for acupuncture points from PubMed, and gathered clinical trial evidence related to acupuncture points respectively. The retrieved data is then formatted into natural language statements to make it more interpretable for the LLM.

**Figure 5.**
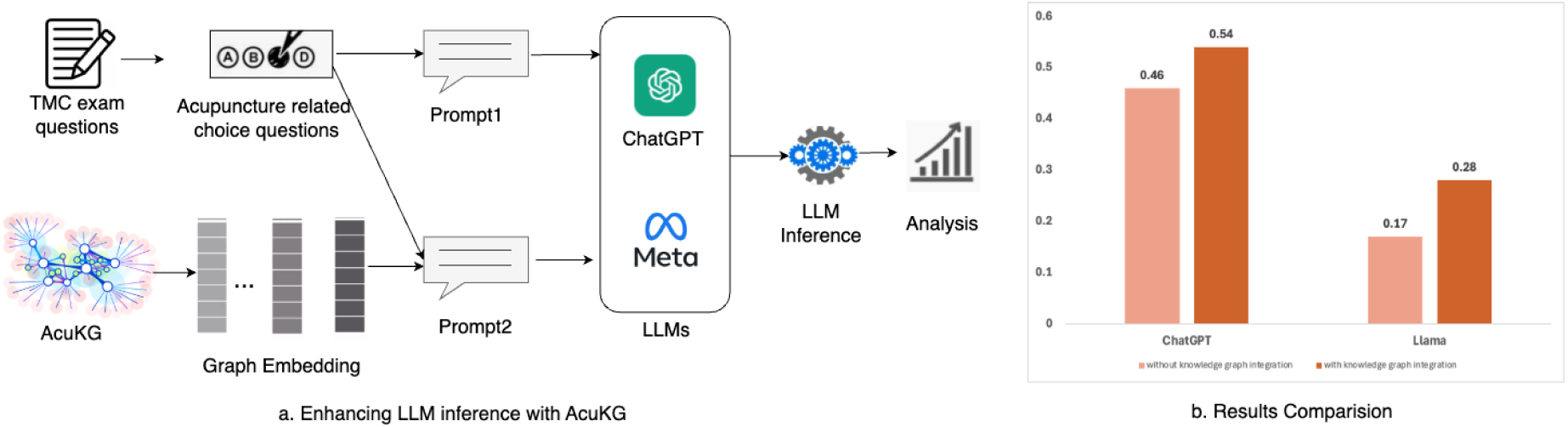
Overview of Enhancing LLM acupuncture-related knowledge accuracy with the integration of AcuKG (a) **Schematic overview** This part illustrates the workflow of integrating AcuKG with LLMs (ChatGPT and LLaMA3) to improve acupuncture-related knowledge accuracy. It shows the initial query process, knowledge graph integration, and enhanced response generation (b) **Results comparison** This part presents accuracy rates of ChatGPT (46% → 54%, P=0.03) and LLaMA3 (17% → 28%, P=0.01) before and after AcuKG integration, showing significant improvements in answer accuracy related to acupoint questions The initial results indicated that while ChatGPT and LLaMA3 were able to generate generally correct answers, its accuracy was limited when it came to specific details. For example, the model struggled with providing precise acupoint locations and sometimes conflated the functions of different acupoints. The accuracy rate of ChatGPT and LLaMA3 in this initial test was approximately 46%, and 17% respectively, highlighting its limitations in this specialized domain.

Once the data is structured, we construct a comprehensive prompt by combining all the formatted KG information with the user’s query. The prompt (provided in supplementary files) is then processed by the large language model, which generates an answer based on both the injected knowledge and its pre-trained linguistic capabilities. By integrating domain-specific knowledge with LLM, we aimed to enhance the model’s accuracy and reliability.

After integrating the acupuncture KG, we re-tested the same questions. To compare the performance before and after the KG integration, we employed the McNemar test, which evaluates the statistical significance of changes in model accuracy. The results showed a significant improvement in the accuracy of ChatGPT and LLaMA3’s responses, with the accuracy rate increasing to approximately 54% (*P*=0.03), and 28% (*P*=0.01) respectively. The model’s ability to provide precise acupoint locations and correct indications for specific conditions was improved. This enhancement suggests that while LLM provides a strong foundation for natural language understanding, its performance on acupuncture-related questions can be significantly enhanced by integrating it with a specialized knowledge graph.

The implications of these findings for medical education are profound. As LLMs like GPT-4o and LLaMA3 continue to advance, they hold great potential as educational tools, particularly when combined with domain-specific resources. In medical education, where accuracy and precision are paramount, the integration of LLMs with specialized knowledge graphs could provide students and practitioners with more reliable and contextually relevant information. This approach could be especially valuable in teaching complex and abstract subjects like acupuncture, where traditional texts and modern clinical knowledge intersect.

Overall, this use case underscores the importance of integrating domain-specific knowledge with LLMs to improve their accuracy and utility in specialized fields. As LLMs become increasingly integrated into medical education and practice, ensuring their reliability and relevance through such enhancements will be crucial for their effective use.

## Discussion

This study focuses on the construction of a comprehensive acupuncture KG using various data sources, emphasizing the integration of structured domain knowledge to enhance the accessibility and utility of acupuncture-related information. The KG captures key relationships between acupoints, anatomical locations, and their therapeutic indications, providing a detailed, queryable framework for representing traditional acupuncture knowledge. The systematic design and development of the KG ensure that complex, interrelated knowledge is organized in a way that is both computationally efficient and clinically meaningful. As a result, our study demonstrated that AcuKG offers enhanced content coverage and denser connections compared to existing acupuncture knowledge graphs. It also enhances the discoverability and application of acupuncture knowledge, supports acupuncture research by providing extensive structured data, and has the potential to advance modern practices through integration with digital devices and electronic acupuncture applications.

The use of multiple data sources—such as web sources, acupuncture textbooks, clinical trials, and biomedical literature—greatly enhances the robustness and comprehensiveness of the acupuncture knowledge graph. Data integration in this study supports the linking of traditional theoretical knowledge with modern clinical and evidence-based insights, ensuring that the graph is both comprehensive and relevant for clinical use. The integration process allowed for cross-referencing information from different domains, improving the graph’s utility in acupuncture knowledge discovery, and enhancing the evidence base for acupuncture practice. For example, clinicians can query the knowledge graph to identify effective acupoints and potential acupoints for specific conditions, and the integration of clinical trials and peer-reviewed literature into the acupuncture knowledge graph ensures that the knowledge discovery is supported by evidence. In addition, the combination of different sources, including web sources, guidelines, with dynamic, real-time data from publications and clinical trials, allows the graph to reflect the evolving landscape of acupuncture research and practice. To this end, AcuKG plays a pivotal role in facilitating knowledge retrieval and knowledge discovery by organizing and presenting acupuncture knowledge in a structured, accessible format.

The primary contributions of this study lie in (1) Introducing a structured knowledge representation for acupuncture: This study develops a graph-based framework to systematically organize acupuncture knowledge, addressing the lack of standardized digital resources in the field. Unlike traditional databases or text-based repositories, AcuKG provides a scalable and flexible platform for encoding and querying complex relations between acupoints, anatomical locations, and therapeutic indications. This structured representation enables a more holistic and interconnected understanding of acupuncture principles. (2) Facilitating efficient knowledge retrieval and complex reasoning: The graph-based approach overcomes challenges posed by fragmented or inaccessible knowledge sources by enabling efficient storage, retrieval, and advanced queries. Researchers and practitioners can easily identify acupoints related to specific conditions, explore relations between adjacent acupoints along meridians, and analyze treatment pathways—tasks that are cumbersome in conventional data storage systems. This structured framework enhances the accessibility and usability of acupuncture knowledge. (3) Enhancing standardization and interoperability with medical ontologies: By integrating widely accepted anatomical ontologies, including UBERON, SNOMED-CT and MeSH, this study ensures consistency in terminology and classification of acupuncture points. Standardization mapping of acupoints to anatomical locations and conditions reduces ambiguity, improves data normalization across sources and enhances interoperability between acupuncture knowledge and broader biomedical datasets, fostering integration with modern medical informatics systems. (4) Improving LLM accuracy and reducing hallucinations through knowledge injection: The study evaluates ChatGPT and Llama using Internal Medicine Question Bank for Chinese Medicine (2020, Acupuncture part), demonstrating that injecting structured acupuncture knowledge significantly enhances model performance in answering expert-level queries. This approach also mitigates a common issue in LLMs—hallucination, where models generate plausible-sounding but factually incorrect responses. By grounding LLM outputs in verifiable, evidence-based knowledge, this study improves the reliability of acupuncture question answering, making LLMs more trustworthy in acupuncture domains, and potentially facilitating AI-driven acupuncture decision support, education, and training.

One limitation of this study is that the relationships extracted from PubMed and ClinicalTrials.gov are co-occurrence relations, as traditional co-occurrence analysis techniques may fail to capture concrete or complex biomedical relations. To address this, future work will focus on developing an advanced NLP pipeline using LLMs, incorporating fine-tuning on domain-specific datasets to enhance the granularity and accuracy of relation extraction from PubMed and ClinicalTrials.gov. Another limitation is that AcuKG is built statically from data sources, preventing automatic updates with newly published studies or clinical trial data. To overcome this, we plan to implement a dynamic synchronization mechanism by integrating automated data ingestion pipelines—such as web scraping for PubMed updates and ClinicalTrials.gov API access—with incremental knowledge graph updating techniques like knowledge graph embeddings and reinforcement learning-based knowledge fusion. This will ensure that AcuKG remains up to date with the latest biomedical findings.

## Conclusion

This study constructs an acupuncture knowledge graph AcuKG by integrating diverse data sources, providing a structured and standard representation of acupuncture knowledge. AcuKG consolidates key information from both acupuncture research and clinical practice, while supporting alignment with established medical ontologies to enhance semantic clarity and interoperability. This work lays the foundation for the development of evidence-based tools in medical acupuncture, contributing to computational framework and knowledge consistency in acupuncture-related information retrieval, reasoning, and clinical application.

## Author contributions

HX, NH and YL designed the study. YL collected, cleaned, and analyzed data from WHO standards and web resources, as well as conducted ontology mapping. XP initially drafted the introduction, analyzed data from PubMed and clinicaltrials.org. DP collected XML data from PubMed and conducted optical character recognition analysis to extract text from WHO Standards in PDF. YH extracted and parsed PubMed data from XML files. NH, SP, and YL designed the use cases. YL conducted the experiments to construct AcuKG and validated the knowledge graph using the designed use cases. SP provided expertise and academic support in Traditional Chinese Medicine. FL provided ontology support. YL conducted statistical analysis. SP and NH contributed to manually reviewing the data. JL provided technical support. YL, NH, QZ, and XP contributed to the visualization results. NH supervised the project. YL drafted the manuscript, while NH, FL, CT and LZ critically reviewed the manuscript.

## Data Availability

The datasets generated and/or analysed during the current study are available in the Github repository, https://github.com/yimingli99/AcuKG-Knowledge-graph-for-medical-acupuncture.

## Reference

[1] Z.-Q. Zhao, “Neural mechanism underlying acupuncture analgesia,” Prog. Neurobiol., vol. 85, no. 4, pp. 355–375, Aug. 2008, doi: 10.1016/j.pneurobio.2008.05.004.

[2] J. Curran, “The Yellow Emperor’s Classic of Internal Medicine,” BMJ, vol. 336, no. 7647, p. 777, Apr. 2008, doi: 10.1136/bmj.39527.472303.4E.

[3] A. White and Editorial Board of Acupuncture in Medicine, “Western medical acupuncture: a definition,” Acupunct. Med. J. Br. Med. Acupunct. Soc., vol. 27, no. 1, pp. 33–35, Mar. 2009, doi: 10.1136/aim.2008.000372.

[4] D. T. Hsu and D. L. Diehl, “Acupuncture. The West gets the point,” Lancet Lond. Engl., vol. 352 Suppl 4, p. SIV1, Dec. 1998, doi: 10.1016/s0140-6736(98)90263-x.

[5] NIH Consensus Conference and M. Bowman, “Acupuncture,” JAMA, vol. 280, no. 17, pp. 1518–1524, Jan. 1998, doi: 10.1001/jama.280.17.1518.

[6] A. Y. Fan, S. H. Stumpf, S. Faggert Alemi, and A. Matecki, “Distribution of licensed acupuncturists and educational institutions in the United States at the start of 2018,” Complement. Ther. Med., vol. 41, pp. 295–301, Dec. 2018, doi: 10.1016/j.ctim.2018.10.015.

[7] T. C. Clarke, L. I. Black, B. J. Stussman, P. M. Barnes, and R. L. Nahin, “Trends in the use of complementary health approaches among adults: United States, 2002-2012,” Natl. Health Stat. Rep., no. 79, pp. 1–16, Feb. 2015.

[8] P. M. Barnes, B. Bloom, and R. L. Nahin, “Complementary and alternative medicine use among adults and children: United States, 2007,” Natl. Health Stat. Rep., no. 12, pp. 1–23, Dec. 2008.

[9] C. Witt et al., “Acupuncture in patients with osteoarthritis of the knee: a randomised trial,” The Lancet, vol. 366, no. 9480, pp. 136–143, Jul. 2005, doi: 10.1016/S0140-6736(05)66871-7.

[10] Y. Li et al., “Development of a Natural Language Processing Tool to Extract Acupuncture Point Location Terms,” in 2023 IEEE 11th International Conference on Healthcare Informatics (ICHI), Jun. 2023, pp. 344–351. doi: 10.1109/ICHI57859.2023.00053.

[11] J. C. Longhurst, “Defining Meridians: A Modern Basis of Understanding,” J. Acupunct. Meridian Stud., vol. 3, no. 2, pp. 67–74, Jun. 2010, doi: 10.1016/S2005-2901(10)60014-3.

[12] Y. Zhao, L. Huang, M. Liu, H. Gao, and W. Li, “Scientific Knowledge Graph of Acupuncture for Migraine: A Bibliometric Analysis from 2000 to 2019,” J. Pain Res., Jun. 2021, Accessed: May 27, 2024. [Online]. Available: https://www.tandfonline.com/doi/abs/10.2147/JPR.S314174

[13] F. Zhang, Y. Shen, H. Fu, H. Zhou, and C. Wang, “Auricular acupuncture for migraine: A systematic review protocol,” Medicine (Baltimore), vol. 99, no. 5, p. e18900, Jan. 2020, doi: 10.1097/MD.0000000000018900.

[14] K. Linde et al., “Acupuncture for Patients With MigraineA Randomized Controlled Trial,” JAMA, vol. 293, no. 17, pp. 2118–2125, May 2005, doi: 10.1001/jama.293.17.2118.

[15] A. J. Vickers et al., “Acupuncture for Chronic Pain: Individual Patient Data Meta-analysis,” Arch. Intern. Med., vol. 172, no. 19, pp. 1444–1453, Oct. 2012, doi: 10.1001/archinternmed.2012.3654.

[16] Y. He et al., “Clinical Evidence for Association of Acupuncture and Acupressure With Improved Cancer Pain,” JAMA Oncol., vol. 6, no. 2, pp. 271–278, Feb. 2020, doi: 10.1001/jamaoncol.2019.5233.

[17] M. Armour et al., “Acupuncture for Depression: A Systematic Review and Meta-Analysis,” J. Clin. Med., vol. 8, no. 8, Art. no. 8, Aug. 2019, doi: 10.3390/jcm8081140.

[18] C. A. Smith, M. Armour, M. S. Lee, L.-Q. Wang, and P. J. Hay, “Acupuncture for depression,” Cochrane Database Syst. Rev., no. 3, 2018, doi: 10.1002/14651858.CD004046.pub4.

[19] G. A. Ulett, J. Han, and S. Han, “Traditional and evidence-based acupuncture,” South Med J, vol. 91, no. 12, Art. no. 12, 1998.

[20] I. Veith, “Acupuncture in traditional Chinese medicine. An historical review.,” Calif. Med., vol. 118, no. 2, Art. no. 2, Feb. 1973.

[21] J. Godwin, “Rising to the Challenges of Evidence-Based Medicine: A Way Forward for Acupuncture,” J. Altern. Complement. Med., vol. 20, no. 11, Art. no. 11, Nov. 2014, doi: 10.1089/acm.2014.0213.

[22] J. J. Hao and M. Mittelman, “Acupuncture: Past, Present, and Future,” Glob. Adv. Health Med., vol. 3, no. 4, pp. 6–8, Jul. 2014, doi: 10.7453/gahmj.2014.042.

[23] W.-H. Kuo, “You’ve got the point?: Acupuncture and the techno-politics of bodyscape,” in Global health and the new world order, Manchester University Press, 2020, pp. 130–153. Accessed: May 28, 2024. [Online]. Available: https://www.manchesterhive.com/display/9781526149688/9781526149688.00011.xml

[24] P. E. Neumann, M. W. Halle, J. Kong, and R. Kikinis, “West meets east: Taking a stab at acupuncture point names,” Clin. Anat., vol. 36, no. 4, pp. 641–650, 2023, doi: 10.1002/ca.24011.

[25] N. Wiseman and K. Boss, Glossary of Chinese Medical Terms and Acupuncture Points. Paradigm Publications, 1990.

[26] D. Gašević, D. Djurić, and V. Devedžić, Eds., “Ontologies,” in *Model Driven Architecture and Ontology Development*, Berlin, Heidelberg: Springer, 2006, pp. 45–77. doi: 10.1007/3-540-32182-9_2.

[27] H. Guan and D. Xie, “Research on building of acupuncture domain ontology,” 2017 IEEE 19th Int. Conf. E-Health Netw. Appl. Serv. Heal., pp. 1–4, Oct. 2017, doi: 10.1109/HealthCom.2017.8210821.

[28] X. Han, X. Li, Y. Liang, X. Wang, D. Xu, and R. Guan, “Acupuncture and Tuina Knowledge Graph for Ancient Literature of Traditional Chinese Medicine,” in 2021 IEEE International Conference on Bioinformatics and Biomedicine (BIBM), Dec. 2021, pp. 674–677. doi: 10.1109/BIBM52615.2021.9669508.

[29] X. Li, X. Han, S. Wei, Y. Liang, and R. Guan, “Acupuncture and tuina knowledge graph with prompt learning,” *Front*. Big Data, vol. 7, Apr. 2024, doi: 10.3389/fdata.2024.1346958.

[30] L. Bos, Medical and Care Compunetics 3. IOS Press, 2006.

[31] M. O’Neil, C. Payne, and J. Read, “Read Codes Version 3: A User Led Terminology,” Methods Inf. Med., vol. 34, no. 1/2, Art. no. 1/2, 1995, doi: 10.1055/s-0038-1634585.

[32] Y. Li et al., “Relation extraction using large language models: a case study on acupuncture point locations,” J. Am. Med. Inform. Assoc., vol. 31, no. 11, Art. no. 11, Nov. 2024, doi: 10.1093/jamia/ocae233.

[33] World Health Organization. Regional Office for the Western Pacific, WHO standard acupuncture point locations in the Western Pacific region. WHO Regional Office for the Western Pacific, 2008. Accessed: Oct. 19, 2022. [Online]. Available: https://apps.who.int/iris/handle/10665/353407

[34] “Find an Acupuncturists, Acupuncture School, Acupuncture Class, News, Articles and Information.” Accessed: May 29, 2024. [Online]. Available: https://www.acufinder.com/

[35] “Home | ClinicalTrials.gov.” Accessed: May 29, 2024. [Online]. Available: https://clinicaltrials.gov/

[36] W. E. Allen, “Terminologia anatomica: international anatomical terminology and Terminologia Histologica: International Terms for Human Cytology and Histology,” J. Anat., vol. 215, no. 2, p. 221, Aug. 2009, doi: 10.1111/j.1469-7580.2009.1093_1.x.

[37] C. J. Mungall, C. Torniai, G. V. Gkoutos, S. E. Lewis, and M. A. Haendel, “Uberon, an integrative multi-species anatomy ontology,” Genome Biol., vol. 13, no. 1, p. R5, Jan. 2012, doi: 10.1186/gb-2012-13-1-r5.

[38] International Health Terminology Standards Development Organisation, “Systematized Nomenclature of Medicine – Clinical Terms (SNOMED CT),” 2019. [Online]. Available: https://www.snomed.org/

[39] US National Library of Medicine, “Medical Subject Headings (MeSH),” 2021. [Online]. Available: https://meshb.nlm.nih.gov/

[40] S. El-Sappagh, F. Franda, F. Ali, and K.-S. Kwak, “SNOMED CT standard ontology based on the ontology for general medical science,” BMC Med. Inform. Decis. Mak., vol. 18, no. 1, Art. no. 1, Aug. 2018, doi: 10.1186/s12911-018-0651-5.

[41] “The Foundations of Chinese Medicine.” Accessed: Mar. 09, 2023. [Online]. Available: https://shop.elsevier.com/books/the-foundations-of-chinese-medicine/maciocia/978-0-7020-5216-3

[42] J. O’Connor and D. Bensky, Acupuncture, A Comprehensive Text. Seattle: Eastland Press, 1981.

[43] Xinnong Cheng, Chinese Acupuncture and Moxibustion. Beijing: Foreign Languages Press, 1987.

[44] “PubMed,” PubMed. Accessed: Dec. 19, 2023. [Online]. Available: https://pubmed.ncbi.nlm.nih.gov/

[45] “yimingli99/AcuKG-Knowledge-graph-for-medical-acupuncture.” Accessed: Mar. 08, 2025. [Online]. Available: https://github.com/yimingli99/AcuKG-Knowledge-graph-for-medical-acupuncture/tree/main

[46] T. Benson, Principles of Health Interoperability HL7 and SNOMED. Springer Science & Business Media, 2012.

[47] “Health Information Technology and Health Data Standards at NLM.” Accessed: May 30, 2024. [Online]. Available: https://www.nlm.nih.gov/healthit/index.html

[48] F. B. Rogers, “Medical subject headings,” Bull. Med. Libr. Assoc., vol. 51, no. 1, pp. 114–116, Jan. 1963.

[49] “Apache PDFBox | A Java PDF Library.” Accessed: Jan. 09, 2025. [Online]. Available: https://pdfbox.apache.org/

[50] “Find an Acupuncturists, Acupuncture School, Acupuncture Class, News, Articles and Information.” Accessed: Jan. 09, 2025. [Online]. Available: https://www.acufinder.com/

[51] “PubMed Central (PMC),” PubMed Central (PMC). Accessed: Mar. 08, 2025. [Online]. Available: https://pmc.ncbi.nlm.nih.gov/

[52] “Neo4j Graph Database & Analytics – The Leader in Graph Databases,” Graph Database & Analytics. Accessed: Mar. 04, 2025. [Online]. Available: https://neo4j.com/

[53] py2neo: Python client library and toolkit for Neo4j. Python. Accessed: Mar. 04, 2025. [OS Independent]. Available: https://py2neo.org/

[54] G. a. Bray, K. k. Kim, J. p. h. Wilding, and on behalf of the W. O. Federation, “Obesity: a chronic relapsing progressive disease process. A position statement of the World Obesity Federation,” Obes. Rev., vol. 18, no. 7, Art. no. 7, 2017, doi: 10.1111/obr.12551.

[55] A. Mukhopadhyay, M. Bhadra, and K. Bose, “Human Obesity: A Background”.

[56] H. J. Lim, H. Xue, and Y. Wang, “Global Trends in Obesity,” in *Handbook of Eating and Drinking: Interdisciplinary Perspectives*, H. L. Meiselman, Ed., Cham: Springer International Publishing, 2020, pp. 1217–1235. doi: 10.1007/978-3-030-14504-0_157.

[57] S. Jalaba, H. Trudeau, and S. Carlson, “Obesity Prevention,” Physician Assist. Clin., vol. 7, no. 1, Art. no. 1, Jan. 2022, doi: 10.1016/j.cpha.2021.07.004.

[58] P. E. Scherer and J. A. Hill, “Obesity, Diabetes, and Cardiovascular Diseases,” Circ. Res., vol. 118, no. 11, Art. no. 11, May 2016, doi: 10.1161/CIRCRESAHA.116.308999.

[59] K. I. Avgerinos, N. Spyrou, C. S. Mantzoros, and M. Dalamaga, “Obesity and cancer risk: Emerging biological mechanisms and perspectives,” Metabolism, vol. 92, pp. 121–135, Mar. 2019, doi: 10.1016/j.metabol.2018.11.001.

[60] Y. Sui et al., “A systematic review on use of Chinese medicine and acupuncture for treatment of obesity,” Aug. 2024, Accessed: Aug. 02, 2024. [Online]. Available: https://onlinelibrary.wiley.com/doi/10.1111/j.1467-789X.2011.00979.x

[61] Y. Li, F. Li, K. Roberts, L. Cui, C. Tao, and H. Xu, “A Comparative Study of Recent Large Language Models on Generating Hospital Discharge Summaries for Lung Cancer Patients,” *arXiv.org*, 2024, doi: 10.48550/arXiv.2411.03805.

[62] C. Tao et al., “VaxBot-HPV: A GPT-based Chatbot for Answering HPV Vaccine-related Questions,” Res. Sq., p. rs.3.rs, Sep. 2024, doi: 10.21203/rs.3.rs-4876692/v1.

[63] J. Li et al., “Mapping vaccine names in clinical trials to vaccine ontology using cascaded fine-tuned domain-specific language models,” J. Biomed. Semant., vol. 15, no. 1, Art. no. 1, Aug. 2024, doi: 10.1186/s13326-024-00318-x.

[64] Y. Li et al., “Improving entity recognition using ensembles of deep learning and fine-tuned large language models: A case study on adverse event extraction from VAERS and social media,” J. Biomed. Inform., vol. 163, p. 104789, Mar. 2025, doi: 10.1016/j.jbi.2025.104789.

[65] Y. Li, J. Li, J. He, and C. Tao, “AE-GPT: Using Large Language Models to extract adverse events from surveillance reports-A use case with influenza vaccine adverse events,” PLOS ONE, vol. 19, no. 3, Art. no. 3, Mar. 2024, doi: 10.1371/journal.pone.0300919.

[66] Y. Li et al., “Artificial intelligence-powered pharmacovigilance: A review of machine and deep learning in clinical text-based adverse drug event detection for benchmark datasets,” J. Biomed. Inform., vol. 152, p. 104621, Apr. 2024, doi: 10.1016/j.jbi.2024.104621.

[67] Y. Hu, et al., “Zero-shot Clinical Entity Recognition using ChatGPT,” *arXiv.org*, 2023, doi: 10.48550/arXiv.2303.16416.

[68] Y. Li et al., “RefAI: a GPT-powered retrieval-augmented generative tool for biomedical literature recommendation and summarization,” J. Am. Med. Inform. Assoc., vol. 31, no. 9, Art. no. 9, Sep. 2024, doi: 10.1093/jamia/ocae129.

[69] Y. Li, Q. Wei, X. Chen, J. Li, C. Tao, and H. Xu, “Improving tabular data extraction in scanned laboratory reports using deep learning models,” J. Biomed. Inform., vol. 159, p. 104735, Nov. 2024, doi: 10.1016/j.jbi.2024.104735.

[70] Y. Li, “Comprehensive Analysis of Adverse Events Following COVID-19 Vaccination: Insights from Diverse Data Sources,” Diss. Theses Open Access, Oct. 2024, [Online]. Available: https://digitalcommons.library.tmc.edu/uthshis_dissertations/66

[71] Y. Li et al., “Enhancing Relation Extraction for COVID-19 Vaccine Shot-Adverse Event Associations with Large Language Models,” Mar. 17, 2025, Research Square. doi: 10.21203/rs.3.rs-6201919/v1.

[72] Y. Li et al., “VaxBot-HPV: a GPT-based chatbot for answering HPV vaccine-related questions,” JAMIA Open, vol. 8, no. 1, p. ooaf005, Feb. 2025, doi: 10.1093/jamiaopen/ooaf005.

[73] A. Gilson et al., “How Does ChatGPT Perform on the United States Medical Licensing Examination (USMLE)? The Implications of Large Language Models for Medical Education and Knowledge Assessment,” JMIR Med. Educ., vol. 9, no. 1, Art. no. 1, Feb. 2023, doi: 10.2196/45312.

[74] T. H. Kung et al., “Performance of ChatGPT on USMLE: Potential for AI-assisted medical education using large language models,” *PLOS Digit*. Health, vol. 2, no. 2, Art. no. 2, Feb. 2023, doi: 10.1371/journal.pdig.0000198.

[75] S. Shahriar et al., “Putting GPT-4o to the Sword: A Comprehensive Evaluation of Language, Vision, Speech, and Multimodal Proficiency,” Preprints, Jun. 2024, doi: 10.20944/preprints202406.1635.v1.

[76] A. Zafar, V. B. Parthasarathy, C. L. Van, S. Shahid, A. I. Khan, and A. Shahid, “Building Trust in Conversational AI: A Review and Solution Architecture Using Large Language Models and Knowledge Graphs,” Big Data Cogn. Comput., vol. 8, no. 6, Art. no. 6, Jun. 2024, doi: 10.3390/bdcc8060070.

